# Implicit provider bias in cancer clinical trial enrollment: A scoping review

**DOI:** 10.1101/2025.06.25.25330296

**Authors:** Melissa Beauchemin, Jessica Verp, Priscila Laforet, Samrawit Solomon, Desiree Walker, Dawn L. Hershman, Edward Bentlyewski, Ni-Cheng Liang, Grace C. Hillyer

## Abstract

**Background:** The lack of racial and ethnic representation in cancer clinical trials further exacerbates existing health disparities and has been linked to inequitable and unequal distribution of resources, social determinants of health, racism, and most recently, provider implicit bias. The purpose of this scoping review is to describe the evidence of implicit bias as it relates to cancer clinical trial enrollment.

**Methods:** Systematic methods were followed to search online databases for publications that assessed provider bias and cancer clinical trial offers through August 1, 2023. Variables of interest included how implicit bias was defined or measured, what type of bias was examined, and if provider implicit bias was identified or described in the cancer clinical trial enrollment process.

**Results:** We identified 10 publications that met the inclusion criteria. Nine of ten studies were conducted in the United States, and all studies utilized observational or qualitative study design. Most assessed the provider perspective, and one study used a validated measure to identify implicit bias. Some studies identified evidence of implicit provider bias in clinical trial discussions and offers; however, it was observed in situations where system constraints, such as lack of resources or time were present that may be common in oncological settings.

**Conclusions:** Multilevel support and dedication to improving racial, ethnic, and other patient-level representativeness are required to mitigate the influence of implicit bias in resource-poor and stressful clinical settings to ensure equitable access to and enrollment in clinical trials for all patients with cancer.

**Registration:** The protocol for this scoping review was registered to Open Science Framework (1), https://osf.io/a6kbt/?view_only=0a857280364a49b9b3f8363b91b00994.

## BACKGROUND

Over the past 10 years, major advancements in the prevention, diagnosis, and treatment of cancer have emerged with the success of immunotherapy, refinement of precision medicine, development of liquid biopsies, and machine learning to make sense of big data (2). However, not all patients with cancer will benefit from these significant and potentially lifesaving medical advances. Only 6.3% of cancer patients participate in clinical trials (3) and, while people who identify from minority racial and ethnic groups comprise 36% of the total U.S. population, fewer than 20% of trial participants are people of color (4). Underrepresentation of participant diversity in clinical trials compromises the generalizability of trial results (5–7), potentially leading to miscalculations of disease-free survival rates and erroneous estimates of treatment efficacy (8) and further exacerbating health disparities (9).

Disparities are the *avoidable* differences that arise from social and economic conditions which determine an individual’s risk of illness, and the actions taken to prevent or treat said illness. Currently, disparities are widely observed in access, treatment, and outcomes between racial and ethnic groups, but they also exist across gender, sexual orientation, age, socioeconomic status, geography, and disability status and, according to Healthy People 2020, “shape an individual’s ability to achieve optimal health” (10). The root causes of health disparities, including the disparity in the representation of racial and ethnic groups in clinical trials, are related to the inequitable and unequal distribution of social, political, economic, and environmental resources and have been linked to the social determinants of health as well as health literacy and structural and systemic racism and other forms of bias (10, 11). One such bias that is recently receiving more attention is provider implicit bias.

Implicit bias is an unconscious form of bias that operates outside of our awareness and may be in direct conflict with one’s espoused values and beliefs (12). It is imprinted on individuals early in life through the process of socialization and surfaces automatically and unintentionally but nonetheless affects judgements, decisions, and behaviors. In contrast, individuals may hold explicit biases (e.g., racism) about which they are very aware and clear (12). Explicit bias can be characterized by overt and extreme negative behaviors (e.g., racial violence, verbal harassment) or acts on a more subtle level such as exclusion (13, 14).

A 2017 systematic review of healthcare professionals displaying implicit bias toward patients concluded that they are not invulnerable to implicit bias and express it at the same level as the general population (15). Provider implicit biases contribute to health disparities by shaping physician behavior resulting in differences in medical treatment based on race, ethnicity, gender, and other patient characteristics (15–17). As a result of exposure to implicit bias, patients may choose not to seek care, not return for recommended follow-up care, or not adhere to treatment plans. This results in higher rates of adverse events and complications of disease and treatment, greater morbidity, and higher mortality rates (15, 18–20).

Demonstration of implicit bias in healthcare exists across all specialties. One study found that physicians are more likely to recommend bypass surgery for White patients than for Black patients. This finding was based on physicians’ belief that Black patients were poorly educated and would not understand the benefit of or take part in post-surgical physical activity for rehabilitation (21). In one review of 77 papers, healthcare providers were found to inadequately treat pain based on patient gender. Reports from women with chronic pain were dismissed due to perceived emotional, hysterical, and sensitive natures, and providers believed that men tended to be more stoic, not seeking pain relief as a risk of appearing weak (22).

Other evidence supports that some physicians withhold recommendations for invasive or aggressive treatments from elderly patients using the rationale that this is a more compassionate choice for the patient (23). It has also been suggested that some physicians delay testing and avoid referrals to specialty care for those with low socioeconomic status to save patients from financial burden (24).

Given evidence supporting that healthcare providers are as likely to exhibit implicit bias as others in the general population and the examples of implicit bias observed throughout healthcare and across specialties, we hypothesize that implicit bias may also influence the enrollment of minority populations to cancer clinical trials. To our knowledge, a scoping review of evidence of implicit bias as it relates to cancer clinical trial enrollment has not been conducted. The purpose of this scoping review is to investigate what is known about provider implicit bias as it relates to clinical decision-making and offers of clinical trial participation to patients who are members of groups underrepresented in cancer clinical trials.

## METHODS

### Review questions

The aim of this scoping review was to determine the value of a future systematic review; summarize and disseminate what is currently known about this topic; and identify gaps in the existing literature. Our objectives included assessing the presence of provider implicit bias in the cancer clinical trial enrollment process; identifying the extent to which implicit bias may be present in the clinical trial enrollment process, if any; to assess how it is being measured; and how it may impact patient/provider clinical trial discussions, and clinical trial recruitment, enrollment, and retention of underrepresented and minority groups with cancer.

### Protocol and registration

This scoping review was developed and written in accordance with the Joanna Briggs Institute (JBI) Manual for Evidence Synthesis and the Preferred Reporting Items for Systematic reviews and Meta-Analyses extension for Scoping Reviews (PRISMA-ScR) (25). The protocol for this scoping review was registered to Open Science Framework (1) and can be found at this link: https://osf.io/a6kbt/?view_only=0a857280364a49b9b3f8363b91b00994

### Inclusion criteria

Journal articles published in English using quantitative and/qualitative methods to measure implicit bias related to cancer-specific prevention, treatment or other types of clinical trials were included in our search. Review papers, commentaries, conference abstracts, or papers not peer-reviewed (e.g., dissertation) were excluded from this scoping review.

<u>Population</u>: For this review, the population of interest included both pediatric and adult cancer patients with a focus on those from underrepresented or minority groups. We chose to include children in this review as evidence suggests that physicians exhibit similar levels of bias in the presence of children and adults (15).

Concept: This review considered papers in which implicit bias as it relates to patient/provider discussions about clinical trials and clinical trial recruitment, enrollment, or retention is reviewed, measured, or analyzed.

Context: Papers considered for this scoping review are within the context of clinical oncology settings in which cancer clinical trial participation might be considered or discussed with patients diagnosed with cancer.

<u>Outcomes</u>: The primary outcomes of interest for this review are the identification of implicit bias in the clinical trial enrollment process and the measurements and qualitative assessments of implicit bias. This includes the results of Implicit Association Test (IAT) (26) and priming tasks as well as any alternate calculations or indications. The secondary outcome of interest is the effect of implicit bias on clinical trial enrollment. The quantification or discussion of this effect varies from paper to paper.

### Search strategy

A comprehensive search of Medline (PubMed), Web of Science, and Embase were conducted. The most recent search was conducted on August 18, 2023. In addition to the electronic database search, we also used Google Scholar for grey literature exploration (27). The search strategy was developed in collaboration with an informationist from within our institutional library system. The full search strategies can be found in the Appendix.

### Study selection

After searching the listed databases, we imported the results into Covidence systematic review software (28). This software was used for de-duplication and screening and houses the final dataset. The screening process occurred in two stages: first, we conducted title and abstract screening, followed by full text screening. The two stages of the screening process were completed by four independent reviewers. In both stages, the reviewers closely followed the established eligibility criteria. In the first stage, the four reviewers screened the search results for relevant titles and abstracts. Each reviewer independently screened papers with 10% overlap in papers screened by more than one reviewer (29). For the second stage, two independent reviewers assessed the full texts of the potentially eligible papers. Conflicts were resolved through a third independent reviewer, or when necessary, a discussion between the four reviewers until a consensus could be reached.

### Data charting and synthesis of the findings

The data extraction template was piloted by four independent reviewers. Any discrepancies between the reviewers were discussed until resolved, and revisions to the template were made accordingly. Data elements that were abstracted included: date of publication, information source, first author, characteristics of participant population, study design (where applicable), outcome measure(s) used (where applicable), and findings.

### Critical appraisal of individual sources of evidence

We conducted a thorough examination of the applied analytical and qualitative methods to assess the evidence of each selected study for quality and validity. We utilized the JBI critical appraisal tools for qualitative (30) and analytical cross-sectional studies (31) to identify potential biases. Reviewers independently appraised all included publications, and discrepancies were resolved during group meetings. Both tools consist of an 8-item checklist in which reviewers are asked yes or no questions with the option to answer unclear or not applicable. The total scores were calculated, and the studies were ranked based on their scores. Next, reviewers sorted the study rankings into three quantiles, low, intermediate, and high quality. The critical appraisal scores informed us of our interpretation and synthesis of study results.

### Synthesis of results

Reviewers relied on the data charting information to summarize and group studies together. Two tables were created to facilitate these discussions: the first summarized individual study characteristics, and the second outlined key findings and implications. Synthesis of findings was discussed and decided upon during sequential team meetings.

## RESULTS AND DISCUSSION

### Study identification

The total combined yield of our search was 5,034 with 3,918 unique papers after de-duplication. There were 103 papers included in the full-text screening, of which 93 were excluding during screening. The most common reasons for exclusion during the full-text stage were publication type [n = 33], wrong outcomes [n = 32], and topic not related to clinical trial enrollment [n = 19]. A total of 10 papers were included in this review (32–41). A PRISMA flowchart summarizing the screening process is displayed in Figure 1.

**Fig. 1:**
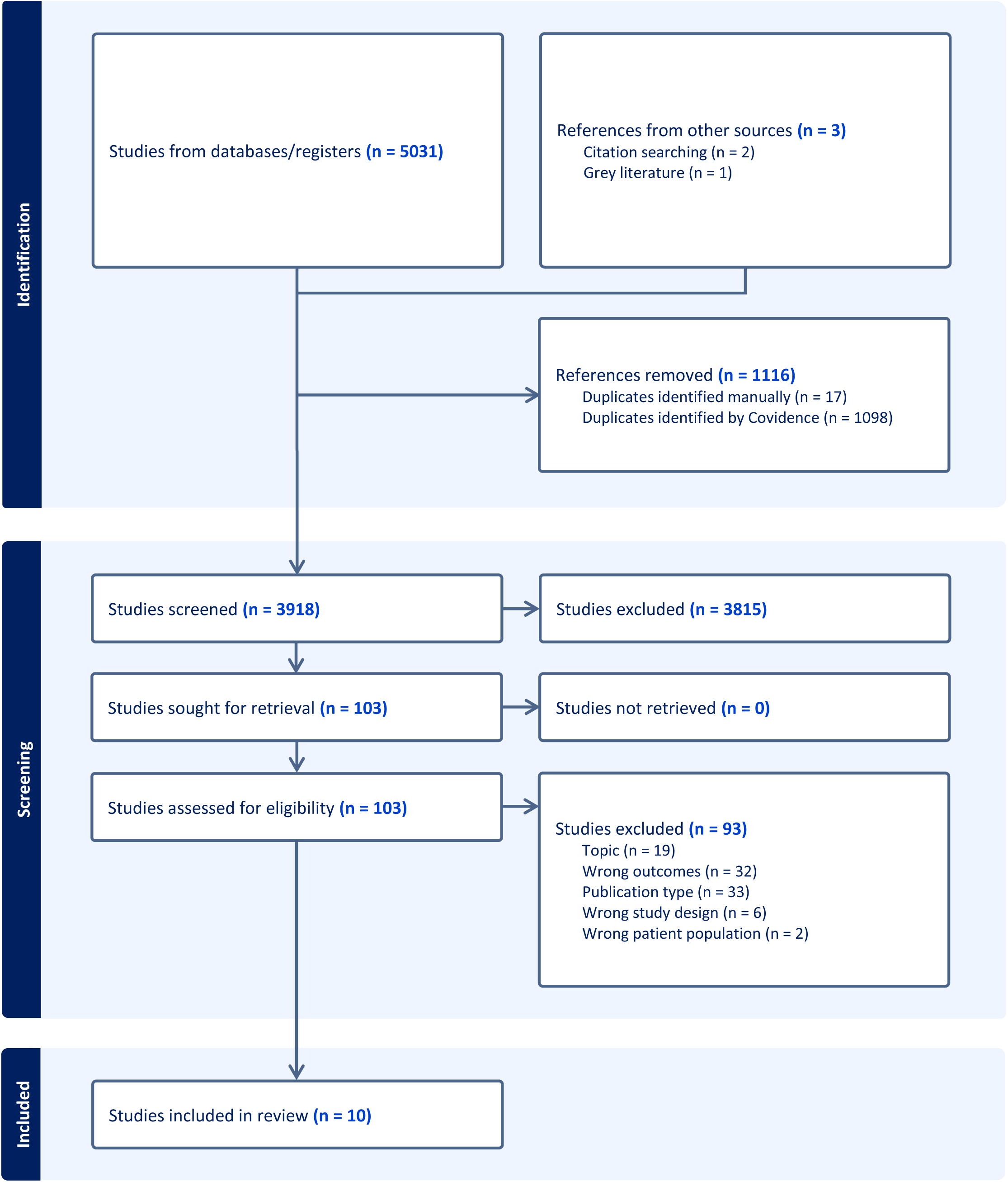
**PRISMA Diagram for Implicit Bias in Clinical Trial Enrollment**

### Characteristics of sources of evidence

The 10 studies included in this scoping review were conducted primarily in the United States (n=9) (32, 33, 35–41), with a single study performed in the Netherlands (34) and all were observational in design using quantitative (32, 33, 35, 37, 40), qualitative (34, 36, 39, 41), or mixed (38) research methods (Table 1). Most were multi-site studies performed in a variety of settings ranging from primary care practices (32) to community (33), academic (34), specialized hospitals (33, 37), and cancer centers (35, 39–41). One study was embedded in the Cancer and Leukemia Group B (CALGB) clinical trial (38) and another was conducted in a non-clinical community center among community members rather than patients (36). Participants included physicians (32–34, 37, 39, 41) (primary care physicians and pediatric, adult, and community oncologists), advanced practice providers (37), research staff (39), cancer patients (35, 40) or parents of cancer patients (33), and healthy community members (36). Bias related to race and/or ethnicity (32, 33, 35–37, 39, 40) was most frequently examined, followed by age (32, 34, 38, 41).

**Table 1.**
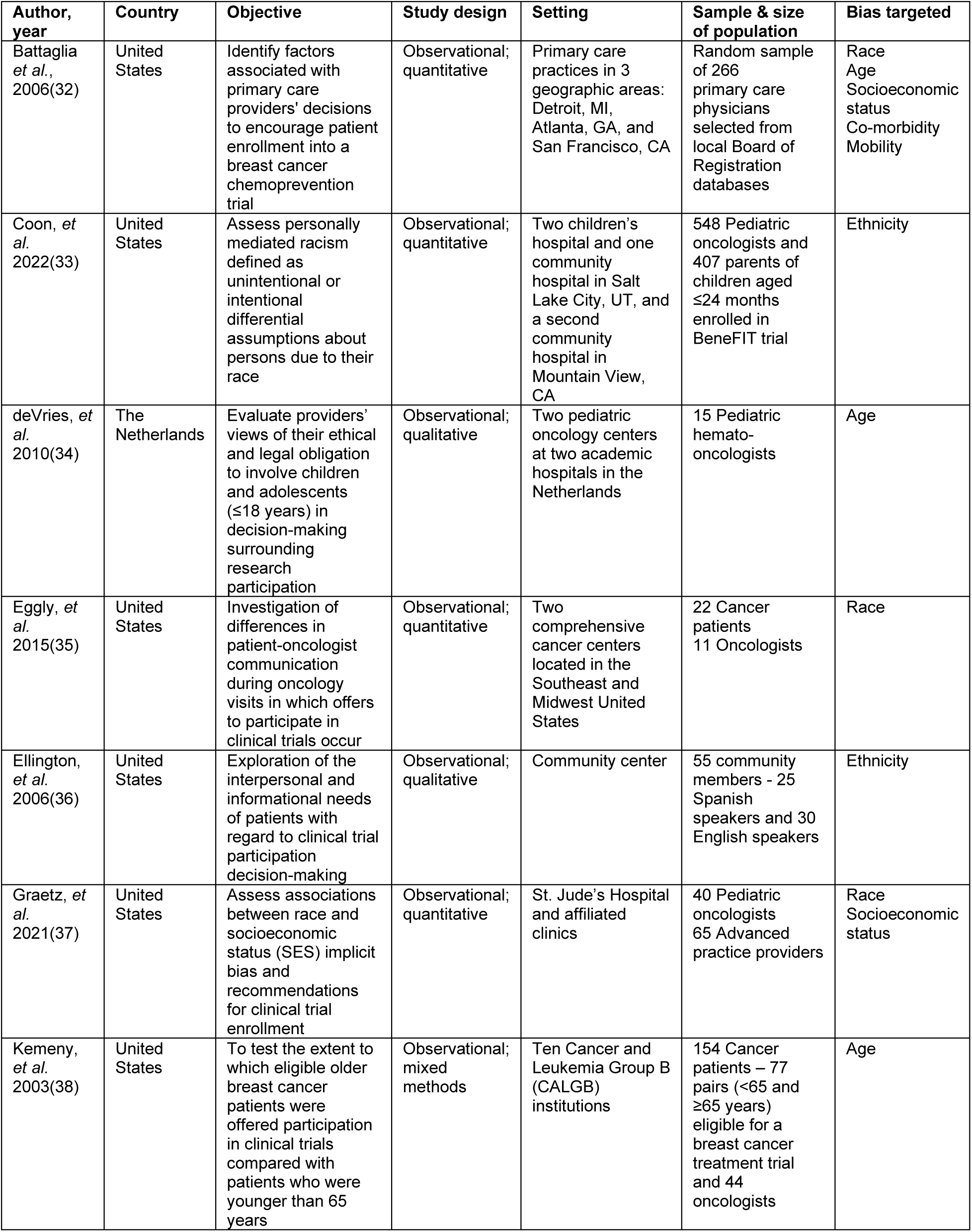
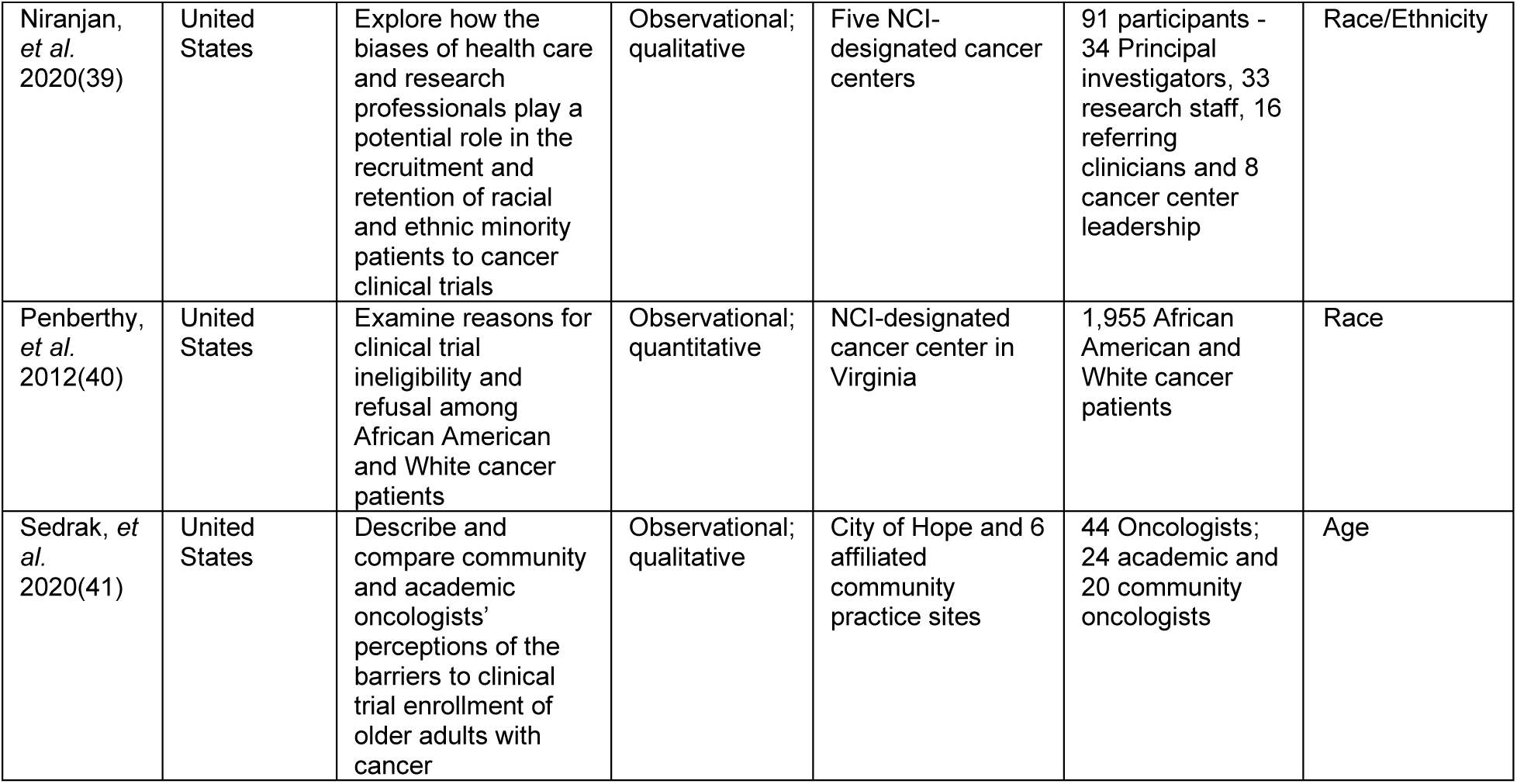
Summary of Research Methods from Studies Examining the Relationship Between Implicit Provider Bias and Cancer Clinical Trial Enrollment.

### Key findings

More than half of the studies (6/10) reflected the perspective of the provider alone (32, 34, 35, 37, 40, 41) (Table 2). To assess potential associations between implicit bias, patient characteristics, and a clinical trial outcome (discussion with participant, clinical trial offer, or enrollment onto clinical trial), a variety of methods were utilized. Five of the 10 studies reviewed used quantitative methods including self-administered surveys (32, 37), evaluation of vignettes (32), and secondary analysis of patient video (35), or study records (33, 40) and four employed qualitative methods such as in-depth or semi-structured interviews (34, 39, 41) and focus groups (36). One study utilized mixed methods that included patient interviews and a provider survey (38). Among studies that used a quantitative measure, only one study directly assessed implicit bias using the IAT (37). The other nine studies sought to identify bias indirectly through observation of communication, behavior, or outcomes.

**Table 2.**
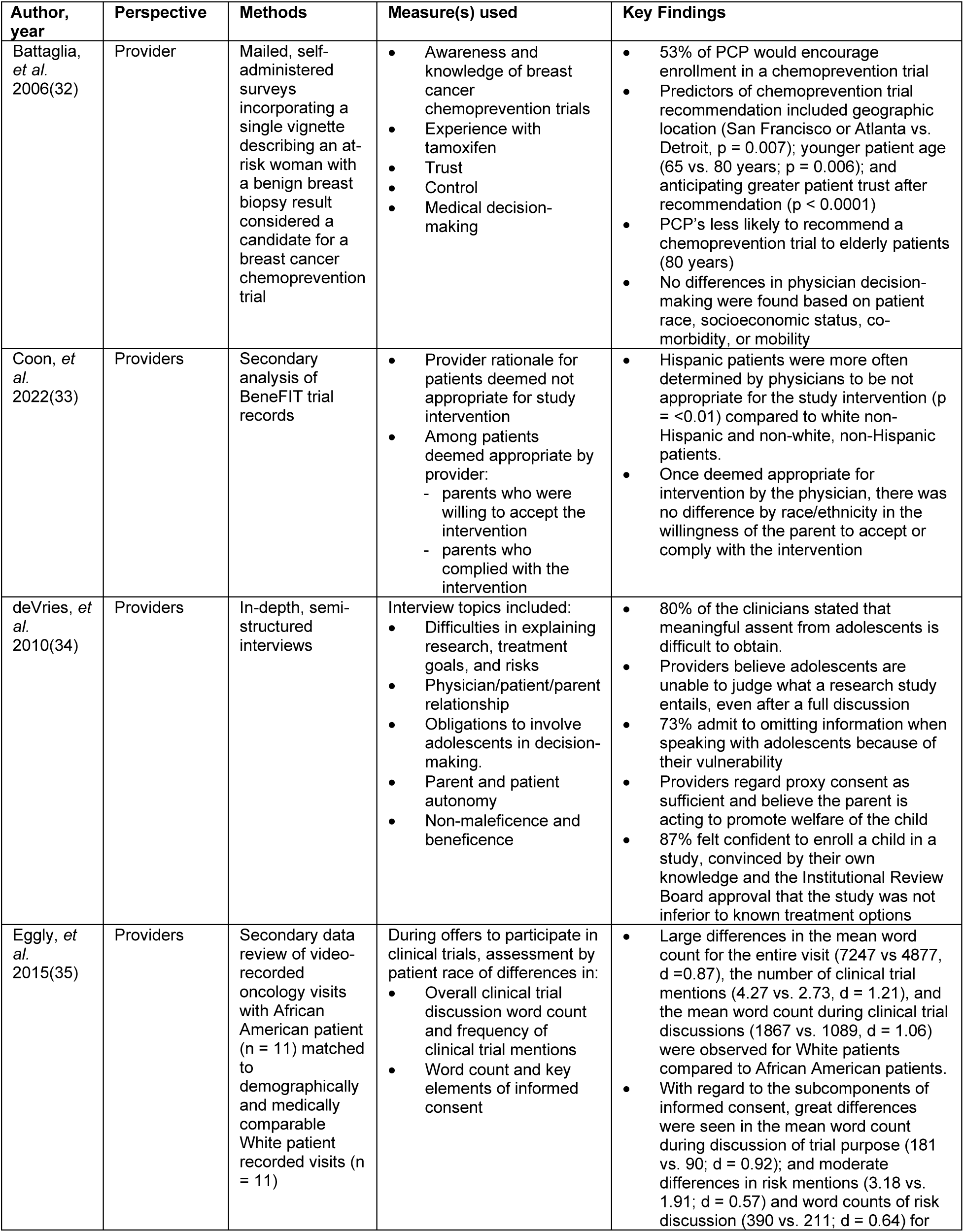
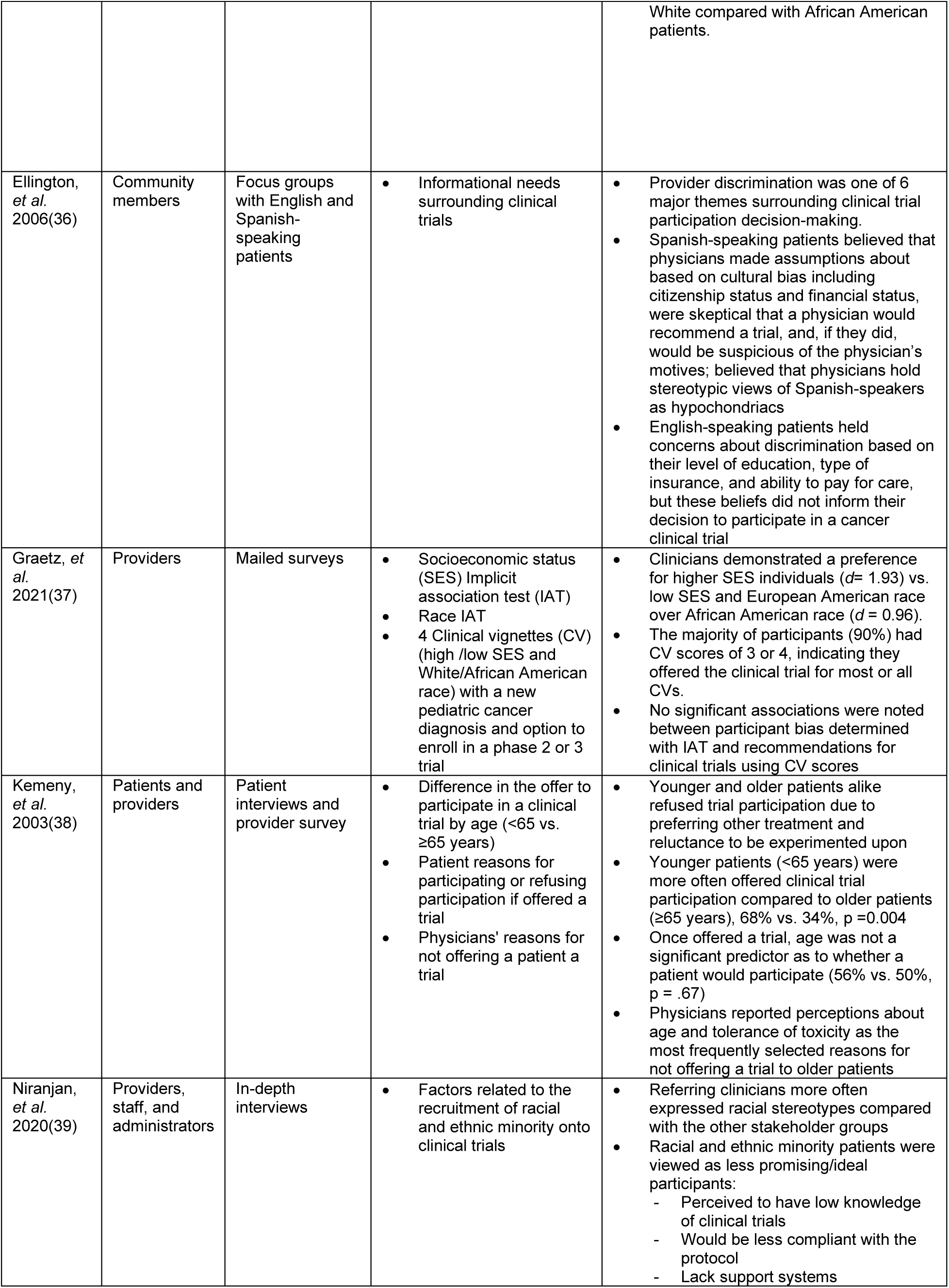
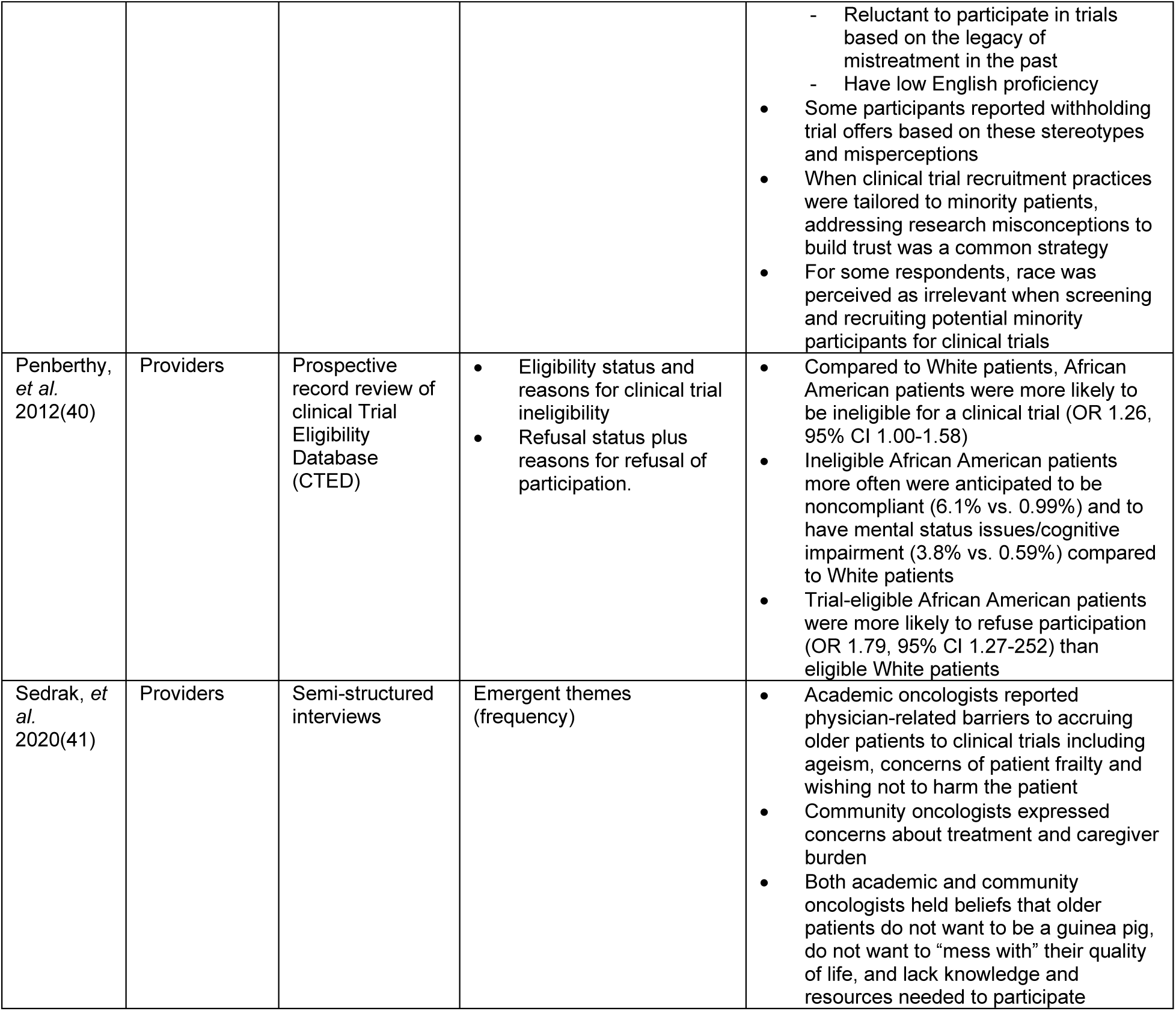
Summary of Key Findings from Studies Examining the Relationship Between Implicit Provider Bias and Cancer Clinical Trial Enrollment.

| Author, year | Perspective | Methods | Measure(s) used | Key Findings |
| --- | --- | --- | --- | --- |
| Battaglia, *et al.* 2006(32) | Provider | Mailed, self-administered surveys incorporating a single vignette describing an at-risk woman with a benign breast biopsy result considered a candidate for a breast cancer chemoprevention trial | Awareness and knowledge of breast cancer chemoprevention trials<br>Experience with tamoxifen<br>Trust<br>Control<br>Medical decision-making | 53% of PCP would encourage enrollment in a chemoprevention trial<br>Predictors of chemoprevention trial recommendation included geographic location (San Francisco or Atlanta vs. Detroit, $p = 0.007$); younger patient age (65 vs. 80 years; $p = 0.006$); and anticipating greater patient trust after recommendation ($p < 0.0001$)<br>PCP's less likely to recommend a chemoprevention trial to elderly patients (80 years)<br>No differences in physician decision-making were found based on patient race, socioeconomic status, co-morbidity, or mobility |
| Coon, *et al.* 2022(33) | Providers | Secondary analysis of BeneFIT trial records | Provider rationale for patients deemed not appropriate for study intervention<br>Among patients deemed appropriate by provider:<br>parents who were willing to accept the intervention<br>parents who complied with the intervention | Hispanic patients were more often determined by physicians to be not appropriate for the study intervention ($p = <0.01$) compared to white non-Hispanic and non-white, non-Hispanic patients.<br>Once deemed appropriate for intervention by the physician, there was no difference by race/ethnicity in the willingness of the parent to accept or comply with the intervention |
| deVries, *et al.* 2010(34) | Providers | In-depth, semi-structured interviews | Interview topics included:<br>Difficulties in explaining research, treatment goals, and risks<br>Physician/patient/parent relationship<br>Obligations to involve adolescents in decision-making.<br>Parent and patient autonomy<br>Non-maleficence and beneficence | 80% of the clinicians stated that meaningful assent from adolescents is difficult to obtain.<br>Providers believe adolescents are unable to judge what a research study entails, even after a full discussion<br>73% admit to omitting information when speaking with adolescents because of their vulnerability<br>Providers regard proxy consent as sufficient and believe the parent is acting to promote welfare of the child<br>87% felt confident to enroll a child in a study, convinced by their own knowledge and the Institutional Review Board approval that the study was not inferior to known treatment options |
| Eggly, *et al.* 2015(35) | Providers | Secondary data review of video-recorded oncology visits with African American patient ($n = 11$) matched to demographically and medically comparable White patient recorded visits ($n = 11$) | During offers to participate in clinical trials, assessment by patient race of differences in:<br>Overall clinical trial discussion word count and frequency of clinical trial mentions<br>Word count and key elements of informed consent | Large differences in the mean word count for the entire visit (7247 vs 4877, $d = 0.87$), the number of clinical trial mentions (4.27 vs. 2.73, $d = 1.21$), and the mean word count during clinical trial discussions (1867 vs. 1089, $d = 1.06$) were observed for White patients compared to African American patients.<br>With regard to the subcomponents of informed consent, great differences were seen in the mean word count during discussion of trial purpose (181 vs. 90; $d = 0.92$); and moderate differences in risk mentions (3.18 vs. 1.91; $d = 0.57$) and word counts of risk discussion (390 vs. 211; $d = 0.64$) for |
|  |  |  |  | White compared with African American patients. |
| Ellington, *et al.* 2006(36) | Community members | Focus groups with English and Spanish-speaking patients | Informational needs surrounding clinical trials | Provider discrimination was one of 6 major themes surrounding clinical trial participation decision-making.<br>Spanish-speaking patients believed that physicians made assumptions about based on cultural bias including citizenship status and financial status, were skeptical that a physician would recommend a trial, and, if they did, would be suspicious of the physician's motives; believed that physicians hold stereotypic views of Spanish-speakers as hypochondriacs<br>English-speaking patients held concerns about discrimination based on their level of education, type of insurance, and ability to pay for care, but these beliefs did not inform their decision to participate in a cancer clinical trial |
| Graetz, *et al.* 2021(37) | Providers | Mailed surveys | Socioeconomic status (SES) Implicit association test (IAT)<br>Race IAT<br>4 Clinical vignettes (CV) (high /low SES and White/African American race) with a new pediatric cancer diagnosis and option to enroll in a phase 2 or 3 trial | Clinicians demonstrated a preference for higher SES individuals ($d = 1.93$) vs. low SES and European American race over African American race ($d = 0.96$).<br>The majority of participants (90%) had CV scores of 3 or 4, indicating they offered the clinical trial for most or all CVs.<br>No significant associations were noted between participant bias determined with IAT and recommendations for clinical trials using CV scores |
| Kemeny, *et al.* 2003(38) | Patients and providers | Patient interviews and provider survey | Difference in the offer to participate in a clinical trial by age (<65 vs. $\geq 65$ years)<br>Patient reasons for participating or refusing participation if offered a trial<br>Physicians' reasons for not offering a patient a trial | Younger and older patients alike refused trial participation due to preferring other treatment and reluctance to be experimented upon<br>Younger patients (<65 years) were more often offered clinical trial participation compared to older patients ($\geq 65$ years), 68% vs. 34%, $p = 0.004$<br>Once offered a trial, age was not a significant predictor as to whether a patient would participate (56% vs. 50%, $p = .67$)<br>Physicians reported perceptions about age and tolerance of toxicity as the most frequently selected reasons for not offering a trial to older patients |
| Niranjan, *et al.* 2020(39) | Providers, staff, and administrators | In-depth interviews | Factors related to the recruitment of racial and ethnic minority onto clinical trials | Referring clinicians more often expressed racial stereotypes compared with the other stakeholder groups<br>Racial and ethnic minority patients were viewed as less promising/ideal participants:<br>Perceived to have low knowledge of clinical trials<br>Would be less compliant with the protocol<br>Lack support systems |
|  |  |  |  | - Reluctant to participate in trials based on the legacy of mistreatment in the past<br>- Have low English proficiency<br>• Some participants reported withholding trial offers based on these stereotypes and misperceptions<br>• When clinical trial recruitment practices were tailored to minority patients, addressing research misconceptions to build trust was a common strategy<br>• For some respondents, race was perceived as irrelevant when screening and recruiting potential minority participants for clinical trials |
| Penberthy, *et al.* 2012(40) | Providers | Prospective record review of clinical Trial Eligibility Database (CTED) | • Eligibility status and reasons for clinical trial ineligibility<br>• Refusal status plus reasons for refusal of participation. | • Compared to White patients, African American patients were more likely to be ineligible for a clinical trial (OR 1.26, 95% CI 1.00-1.58)<br>• Ineligible African American patients more often were anticipated to be noncompliant (6.1% vs. 0.99%) and to have mental status issues/cognitive impairment (3.8% vs. 0.59%) compared to White patients<br>• Trial-eligible African American patients were more likely to refuse participation (OR 1.79, 95% CI 1.27-252) than eligible White patients |
| Sedrak, *et al.* 2020(41) | Providers | Semi-structured interviews | Emergent themes (frequency) | • Academic oncologists reported physician-related barriers to accruing older patients to clinical trials including ageism, concerns of patient frailty and wishing not to harm the patient<br>• Community oncologists expressed concerns about treatment and caregiver burden<br>• Both academic and community oncologists held beliefs that older patients do not want to be a guinea pig, do not want to “mess with” their quality of life, and lack knowledge and resources needed to participate |

### Findings supporting or refuting the role of implicit bias in cancer clinical trial discussions

The role of implicit bias was not clearly defined across included studies in affecting cancer clinical trial recruitment, discussion, or enrollment. Some studies identified that providers considered racial and ethnic minority patients to be nonideal candidates for participation (33, 39, 40). Niranjan et al. found that not only do physicians find patients from minority racial and ethnic groups less desirable as trial candidates because they are thought to be less compliant, lack support systems, are reluctant to participate due to a legacy of medical mistreatment in the patient and have low English proficiency, but were more inclined to withhold trial offers based on these attitudes (39). Eggly et al. evaluated word counts in clinical trial discussions and the informed consent process and found that, in general trial discussions, word counts were lower in conversations with African American patients compared with White patients as were the word counts in the key components of informed consent including purpose and risk (35).

However, the impact of these perceptions was unclear. Despite a strong preference of physicians for European Americans over African Americans, Graetz et al. found that this racial preference did not impact clinical decision-making with regard to trial participation (37).

Battaglia et al. found no evidence of difference in clinical decision-making related to patient race, socioeconomic status, comorbidity or mobility but did observe disparities in recommendations to participate in a chemoprevention trial based on of the age of the patient among primary care providers (32).

In addition to bias toward certain racial and ethnic patients, other included studies examined the role of age in clinical trial offers. In a qualitative study, DeVries et al. describe pediatric oncologists’ perceptions that adolescents are unable to participate in trial discussions because of their vulnerability; thus, information may be withheld (34). Older age was also identified as a potential factor impacting clinical trial offers, due to concerns about toxicity (38, 41), frailty, treatment burden and perceptions that older patients do not want to be experimented upon or have their quality of life compromised (41).

Kemeny et al. examined the perspectives of physicians and older patients with breast cancer, questioning patients about their reasons for trial refusal (38). Ellington et al. spoke with English and Spanish speaking members of the community and found that community members who identified as Hispanic reported that physicians make assumptions about them because of their cultural background (36). These community members were also skeptical that a doctor would recommend a trial for them because of their stereotypical attitudes toward them. English-speaking patients similarly felt that doctors made assumptions about them, but the assumptions were related to the individual’s socioeconomic status and education rather than culture.

### Critical appraisal within sources of evidence

Of the 10 included studies, four studies used qualitative methods and were assessed using the Checklist for Qualitative Research. The remaining six used quantitative methods and were assessed using the JBI Critical Appraisal Checklist for Cross-sectional Research. The qualitative studies proved strong when appraised, with three studies scoring “yes” on 6 of the 8 items. One study scored “yes” on 5 of the 8 items. Three of the four studies fell short of locating the researcher culturally or theoretically in the research and did not describe the influence of the researcher on the research and vice versa. Comparatively, the six quantitative studies fell short. Four of the six studies failed to identify confounding factors. One study had inclusion criteria and outcomes measures deemed “unclear”. Two of the six studies did not clearly state the validity or reliability of the exposure measurement used in the study.

We conducted a robust, thorough review of the literature on the role of implicit provider bias on cancer clinical trial discussions, offers, and enrollment outcomes. Of the ten studies that met our inclusion criteria, we found data to support that the expression of implicit bias in provider behavior may occur; however, the process is multifactorial and complex, involving patient, provider, and systems that influence the potential for discussions of, consent for, and enrollment to cancer clinical trials.

This review provides support that, similar to other healthcare settings, implicit bias may play a role in positive outcomes related to cancer clinical trials. Provider implicit bias is more likely to be activated in situations when the provider is exhausted, very busy, working in stressful and high-pressure conditions, balancing a high cognitive load, and when decisions need to be made when only incomplete or ambiguous information is available (16, 19, 42, 43). These situational factors are commonly experienced in the oncology setting and were mentioned in the limited number of studies included in this review as heavily influencing provider decisions to discuss or recommend clinical trials.

The measures used across the included studies in this review varied widely with no consistency across studies. Objective measures of implicit bias, such as the IAT, have been shown to be useful in other settings; however, there are also concerns that this and similar measures are limited in their ability to control for situational factors that may trigger biased behaviors, such as measurable differences in discussing cancer clinical trials. In this review, only one study by Graetz et al. used the IAT to measure implicit bias, and though providers preferred individuals from higher socioeconomic status or American European race, the impact of their implicit bias was not associated with clinical trial discussions or offers (37). This study, however, was conducted in a pediatric oncology setting, where participation rates are significantly higher than those observed among adults (44). In contrast to a standardized measure, the study by Eggly et al. (35) used mixed methods to investigate communication during video-recorded oncology visits, identifying a statistically significant difference by patient race in the total word count and the frequency of clinical trial mentions. Future studies may need to balance this time- and resource-intensive approach with a more quantifiable measure that demonstrates validity in comparison.

As with other studies that examined barriers to clinical trial participation, system- and institutional-barriers cannot be ignored as they support or refute the situational factors that may proceed a clinical trial discussion and subsequent enrollment. The qualitative studies, in particular, highlight that it is difficult to consider only one level of barriers (e.g. provider implicit bias) in isolation from the other interrelated levels (patient, institutional, system) (34, 36, 39, 41). This review provides additional support in favor of multilevel strategies to reduce barriers to clinical trial participation, in particular among historically underrepresented populations (6).

Efforts to improve patient-provider trust and communication, simultaneously supporting the resources needed to offer, enrolling, and following patients on cancer clinical trials offer the most promise (45); however, this requires stakeholder engagement and policy-level support. As others have stated, though substantial efforts are currently underway to address barriers to clinical trial participation, substantial changes in oncology culture will require major investments and resources directed at multiple levels in tandem to support equitable and diverse representation in cancer clinical trials (46).

### Limitations

We followed established methodology to conduct and synthesize this integrative review; however, there are limitations to this study. It is possible that our search strategy, though exhaustive, may not have included all possible terms for implicit bias. Further, publication bias is a possible concern because studies with negative findings or limited statistical significance may not be searchable even using grey literature search strategies. Our study is strengthened by our rigorous methodological approach and team-based review process; we also have representation across multiple disciplines and patient perspectives.

## CONCLUSIONS

We present a comprehensive literature review on the impact of implicit provider bias as it relates to discussions, offers, and enrollment outcomes in cancer clinical trials. Our findings indicate that implicit bias may influence the enrollment process but also demonstrate the complexity of enrolling patients to clinical trials, involving factors related to patients, providers, and systems. Our study highlights that situational factors such as provider exhaustion and high-pressure conditions can activate implicit bias, especially in the oncology setting. The studies included varied in measurement methods, and while the IAT was used in one study, its impact on clinical trial discussions was inconclusive creating a challenge in assessing the first and most critical step in the clinical trial enrollment process. Our findings emphasize the need for multilevel strategies to address barriers to clinical trial participation, including patient-provider trust, communication, and systemic support. However, it also acknowledges the challenges of implementing substantial changes in oncology culture, requiring significant investments and policy-level support.

## Abbreviations

CALGB: Cancer and Leukemia Group B
IAT: Implicit Association Test
JBI: Joanna Briggs Institute
PRISMA-ScR: Preferred Reporting Items for Systematic reviews and Meta-Analyses extension for Scoping Reviews
US: United States.

## Declarations

### Ethics approval

Not applicable.

### Consent for publication

Not applicable.

### Availability of data and materials

The database supporting the conclusions of this article is available in the Open Science Framework (1), https://osf.io/a6kbt/?view_only=0a857280364a49b9b3f8363b91b00994

### Authors’ contributions

MB and GCH were responsible for the conceptualization, methodology, validations, formal analysis, and writing of the original and final drafts of the paper. MB supervised research activity planning and execution. GCH acquired financial support for the project. JV, PL, and SS curated the data, prepared the data presentation, and contributed to the writing of the original and final drafts. DW, DLH, EB, and N-CL provided critical review, commentary, and revision of the final draft.

## Data Availability

The database supporting the conclusions of this article is available in the Open Science Framework

https://osf.io/a6kbt/?view_only=0a857280364a49b9b3f8363b91b00994

## Acknowledgements

Not applicable.

## Appendix

**PICO Statement**

|  |  |
| --- | --- |
| **P** | ("neoplas*" [tiab] OR "tumor*" [tiab] OR "tumour*" [tiab] OR "cancer*" [tiab] OR "malignan*" [tiab] OR "oncolog*" [tiab]) |
| **C** | ("implicit association test*" [tiab] OR "IAT" [tiab] OR "implicit attitude*" [tiab] OR ("implicit" AND "bias*") OR "implicit bias*" [tiab] OR (prejudice AND implicit) OR "unconscious bias*" [tiab] OR "cognitive bias*" [tiab] OR "implicit preference*" [tiab] OR "sequential priming task*" [tiab] OR "stereotype priming" [tiab] OR "priming task*" [tiab] OR "stereotyp*" [tiab] OR "implicit prejudice*" [tiab] OR "attitude*" [tiab]) |
| **C** | ("trial*" [tiab] OR "clinical research" [tiab] OR "clinical research trial*" [tiab] OR "clinical stud*" [tiab] OR "clinical trial*" [tiab]) |

## REFERENCES

1. Center for Open Science. Open Science Framework. 2024. https://osf.io/. Accessed 6 Feb 2024.

2. McDowell S, Ludwig Rausch S, Simmons K. Cancer research insights from the latest decade, 2010 to 2020. 2019. https://www.cancer.org/research/acs-research-news/cancer-research-insights-from-the-latest-decade-2010-to-2020.html. Accessed 18 Aug 2023.

3. Unger JM, Fleury M. Nationally representative estimates of the participation of cancer patients in clinical research studies according to the commission on cancer. J Clin Oncol. 2021;39 Suppl 28:74–74.

4. Melillo G. Racial bias may impact minority participation in cancer trials. 2020. https://www.ajmc.com/view/racial-bias-may-impact-minority-participation-in-cancer-trials. Accessed 18 Aug 2023.

5. Ford JG, Howerton MW, Lai GY, Gary TL, Bolen S, Gibbons MC, et al. Barriers to recruiting underrepresented populations to cancer clinical trials: a systematic review. Cancer. 2008;112:228–42.

6. Hamel LM, Penner LA, Albrecht TL, Heath E, Gwede CK, Eggly S. Barriers to clinical trial enrollment in racial and ethnic minority patients with cancer. Cancer Control. 2016;23:327–37.

7. Kwiatkowski K, Coe K, Bailar JC, Swanson GM. Inclusion of minorities and women in cancer clinical trials, a decade later: have we improved? Cancer. 2013;119:2956–63.

8. Antman K, Amato D, Wood W, Carson J, Suit H, Proppe K, et al. Selection bias in clinical trials. J Clin Oncol. 1985;3:1142–7.

9. Smedley BD, Stith AY, Nelson AR, editors. Unequal treatment: confronting racial and ethnic disparities in healthcare. Washington, DC: National Academy Press; 2003.

10. Healthy People 2030. Health equity and health disparities environmental scan. 2022. https://health.gov/sites/default/files/2022-04/HP2030-HealthEquityEnvironmentalScan.pdf. Accessed 18 Aug 2023.

11. Healthy People 2030. Social determinants of health. 2022. https://health.gov/healthypeople/priority-areas/social-determinants-health. Accessed 18 Aug 2023.

12. National Center for Cultural Competence. Two types of bias. n.d. https://nccc.georgetown.edu/bias/module-3/1.php. Accessed 20 May 2023.

13. Amodio DM, Ratner KG. A memory systems model of implicit social cognition. Current Directions in Psychological Science. 2011;20:143–8.

14. Smith ER, DeCoster J. Dual-process models in social and cognitive psychology: conceptual integration and links to underlying memory systems. Pers Soc Psychol Rev. 2000;4:108–31.

15. FitzGerald C, Hurst S. Implicit bias in healthcare professionals: a systematic review. BMC Med Ethics. 2017;doi:10.1186/s12910-017-0179-8.

16. Alspach JG. Implicit bias in patient care: An endemic blight on quality care. Critical Care Nurse. 2018;38:12–6.

17. Chapman EN, Kaatz A, Carnes M. Physicians and implicit bias: how doctors may unwittingly perpetuate health care disparities. J Gen Intern Med. 2013;28:1504–10.

18. Alspach JG. Because women’s lives matter, we need to eliminate gender bias. Critical Care Nurse. 2017;37:10–8.

19. Burgess DJ. Are providers more likely to contribute to healthcare disparities under high levels of cognitive load? How features of the healthcare setting may lead to biases in medical decision making. Medical Decision Making. 2010;30:246–57.

20. Hall WJ, Chapman MV, Lee KM, Merino YM, Thomas TW, Payne BK, et al. Implicit racial/ethnic bias among health care professionals and its influence on health care outcomes: a systematic review. Am J Public Health. 2015;105:e60–e76.

21. Dovidio JF, Eggly S, Albrecht TL, Hagiwara N, Penner LA. Racial biases in medicine and healthcare disparities. Testing, Psychometrics, Methodology in Applied Psychology. 2016;23:489–510.

22. Samulowitz A, Gremyr I, Eriksson E, Hensing G. “Brave men” and “emotional women“: A theory-guided literature review on gender bias in health care and gendered norms towards patients with chronic pain. Pain Res Manag. 2018;doi:10.1155/2018/6358624.

23. Ben-Harush A, Shiovitz-Ezra S, Doron I, Alon S, Leibovitz A, Golander H, et al. Ageism among physicians, nurses, and social workers: findings from a qualitative study. Eur J Ageing. 2017;14:39–48.

24. Arpey NC, Gaglioti AH, Rosenbaum ME. How socioeconomic status affects patient perceptions of health care: a qualitative study. J Prim Care Community Health. 2017;8:169–75.

25. Aromataris E, Lockwood C, Porritt K, Pilla B, Jordan Z, editors. JBI Manual for Evidence Synthesis: JBI; 2024. https://synthesismanual.jbi.global. 10.46658/JBIMES-20-01. Accessed 14 Jul 2024.

26. Greenwald AG, McGhee DE, Schwartz JLK. Measuring individual differences in implicit cognition: the implicit association test. J Pers Soc Psychol.1998;74:1464–80.

27. Paez A. Gray literature: An important resource in systematic reviews. J Evid Based Med. 2017;10:233–40.

28. Veritas Health Innovation. Covidence systematic review software, Melbourne, Australian. n.d. https://www.covidence.org/. Accessed 6 Feb 2024.

29. Stoll CRT, Izadi S, Fowler S, Green P, Suls J, Colditz GA. The value of a second reviewer for study selection in systematic reviews. Res Synth Methods. 2019;10:539–45.

30. Hannes K, Lockwood C. Pragmatism as the philosophical foundation for the Joanna Briggs meta-aggregative approach to qualitative evidence synthesis. J Adv Nurs. 2011;67:1632–42.

31. Vardell E, Malloy M. Joanna Briggs Institute: an evidence-based practice database. Med Ref Serv Q. 2013;32:434–42.

32. Battaglia TA, Ash A, Prout MN, Freund KM. Cancer prevention trials and primary care physicians: factors associated with recommending trial enrollment. Cancer Detect Prev. 2006;30:34–7.

33. Coon ER, Schroeder AR, Lion KC, Ray KN. Disparities by ethnicity in enrollment of a clinical trial. Pediatrics. 2022;149(2):e2021052595.

34. de Vries MC, Wit JM, Engberts DP, Kaspers GJL, van Leeuwen E. Pediatric oncologists’ attitudes towards involving adolescents in decision-making concerning research participation. Pediatr Blood Cancer. 2010;55:123–8.

35. Eggly S, Barton E, Winckles A, Penner LA, Albrecht TL. A disparity of words: racial differences in oncologist-patient communication about clinical trials. Health Expect. 2015;18:1316–26.

36. Ellington L, Wahab S, Sahami Martin S, Field R, Mooney KH. Factors that influence Spanish- and English-speaking participants’ decision to enroll in cancer randomized clinical trials. Psycho-oncology. 2006;15:273–84.

37. Graetz DE, Madni A, Gossett J, Kang G, Sabin JA, Santana VM, et al. Role of implicit bias in pediatric cancer clinical trials and enrollment recommendations among pediatric oncology providers. Cancer. 2021;127:284–90.

38. Kemeny MM, Peterson BL, Kornblith AB, Muss HB, Wheeler J, Levine E, et al. Barriers to clinical trial participation by older women with breast cancer. J Clin Oncol. 2003;21:2268–75.

39. Niranjan SJ, Martin MY, Fouad MN, Vickers SM, Wenzel JA, Cook ED, et al. Bias and stereotyping among research and clinical professionals: Perspectives on minority recruitment for oncology clinical trials. Cancer. 2020;126:1958–68.

40. Penberthy L, Brown R, Wilson-Genderson M, Dahman B, Ginder G, Siminoff LA. Barriers to therapeutic clinical trials enrollment: differences between African-American and white cancer patients identified at the time of eligibility assessment. Clinical Trials. 2012;9:788–97.

41. Sedrak MS, Mohile SG, Sun V, Sun CL, Chen BT, Li D, et al. Barriers to clinical trial enrollment of older adults with cancer: a qualitative study of the perceptions of community and academic oncologists. J Geriatr Oncol. 2020;11:327–34.

42. Yzerbyt VY, Rogier A, Fiske ST. Group entitativity and social attribution: on translating situational constraints into stereotypes. Pers Soc Psychol Bull. 1998;24:1089–103.

43. Bertrand M, Chugh D, Mullainathan S. Implicit discrimination. American Economic Review. 2005;95:94–8.

44. Faulk KE, Anderson-Mellies A, Cockburn M, Green AL. Assessment of enrollment characteristics for Children’s Oncology Group (COG) upfront therapeutic clinical trials 2004-2015. PloS One. 2020;15:e0230824.

45. Unger JM, Hershman DL, Till C, Minasian LM, Osarogiagbon RU, Fleury ME, et al. “When offered to participate”: a systematic review and meta-analysis of patient agreement to participate in cancer clinical trials. JNCI. 2020;113:244–57.

46. Acuña-Villaorduña A, Baranda JC, Boehmer J, Fashoyin-Aje L, Gore SD. Equitable access to clinical trials: how do we achieve it? American Society of Clinical Oncology Educational Book. 2023(43):e389838.

